# Uncovering clinical risk factors and prediction of severe COVID-19: A machine learning approach based on UK Biobank data

**DOI:** 10.1101/2020.09.18.20197319

**Authors:** Kenneth C.Y. Wong, Yong Xiang, Hon-Cheong So

**Affiliations:** School of Biomedical Sciences, The Chinese University of Hong Kong, Shatin, Hong Kong; CUHK Shenzhen Research Institute, Shenzhen, China; KIZ-CUHK Joint Laboratory of Bioresources and Molecular Research of Common Diseases, Kunming Institute of Zoology and The Chinese University of Hong Kong, China; Department of Psychiatry, The Chinese University of Hong Kong, Shatin, Hong Kong; Margaret K.L. Cheung Research Centre for Management of Parkinsonism, The Chinese University of Hong Kong, Shatin, Hong Kong; Brain and Mind Institute, The Chinese University of Hong Kong, Shatin, Hong Kong; Hong Kong Branch of the Chinese Academy of Sciences Center for Excellence in Animal Evolution and Genetics, The Chinese University of Hong Kong, Hong Kong SAR, China

## Abstract

**Background:** COVID-19 is a major public health concern. Given the extent of the pandemic, it is urgent to identify risk factors associated with disease severity. Accurate prediction of those at risk of developing severe infections is also of high clinical importance.

**Methods:** Based on the UK Biobank(UKBB data), we built machine learning(ML) models to predict the risk of developing severe or fatal infections, and to evaluate major risk factors involved. We first restricted the analysis to infected subjects(*N*=7846), then performed analysis at a population level, considering those with no known infection as controls(*N* for controls=465,728). Hospitalization was used as a proxy for severity. Totally 97 clinical variables(collected prior to COVID-19 outbreak) covering demographic variables, comorbidities, blood measurements(e.g. hematological/liver/renal function/metabolic parameters etc.), anthropometric measures and other risk factors (e.g. smoking/drinking habits) were included as predictors. We also constructed a simplified (‘lite’) prediction model using 27 covariates that can be more easily obtained (demographic and comorbidity data). XGboost (gradient boosted trees) was used for prediction and predictive performance was assessed by cross-validation. Variable importance was quantified by Shapley values and accuracy gain. Shapley dependency and interaction plots were used to evaluate the pattern of relationship between risk factors and outcomes.

**Results:** A total of 2386 severe and 477 fatal cases were identified. For the analysis among infected individuals (*N*=7846),our prediction model achieved AUCs of 0.723(95% CI:0.711-0.736) and 0.814(CI: 0.791-0.838) for severe and fatal infections respectively. The top five contributing factors for severity were age, number of drugs taken(cnt_tx), cystatin C(reflecting renal function), wait-hip ratio (WHR) and Townsend Deprivation index (TDI). For prediction of mortality, the top features were age, testosterone, cnt_tx, waist circumference(WC) and red cell distribution width (RDW).

In analyses involving the whole UKBB population, the corresponding AUCs for severity and fatality were 0.696(CI:0.684-0.708) and 0.802(CI:0.778-0.826) respectively. The same top five risk factors were identified for both outcomes, namely age, cnt_tx, WC, WHR and TDI. Apart from the above features, Type 2 diabetes(T2DM), HbA1c and apolipoprotein A were ranked among the top 10 in at least two (out of four) analyses. Age, cystatin C, TDI and cnt_tx were among the top 10 across all four analyses.

As for the ‘lite’ models, the predictive performances in terms of AUC are broadly similar, with estimated AUCs of 0.716, 0.818, 0.696 and 0.811 respectively. The top-ranked variables were similar to above, including for example age, cnt_tx, WC, male and T2DM.

**Conclusions:** We identified a number of baseline clinical risk factors for severe/fatal infection by an ML approach. For example, age, central obesity, impaired renal function, multi-comorbidities and cardiometabolic abnormalities may predispose to poorer outcomes. The presented prediction models may be useful at a population level to help identify those susceptible to developing severe/fatal infections, hence facilitating targeted prevention strategies. Further replications in independent cohorts are required to verify our findings.

## Introduction

Coronavirus Disease 2019 (COVID-19) has resulted in a pandemic affecting more than a hundred countries worldwide ^1-3^. More than 123 million confirmed cases and 2.7 million fatalities have been reported worldwide as at 22^nd^ Mar 2020 (https://coronavirus.jhu.edu/map.html), while a large number of mild or asymptomatic cases may remain undetected. Given the extent of the pandemic, it is urgent to identify risk factors that may be associated with severe disease, and to gain deeper understanding its pathophysiology. Accurate prediction of those at risk of developing severe infections is also clinically very important.

Machine learning (ML) approaches are powerful tools to predict disease outcomes and have been increasingly applied in biomedical research. An important advantage is that ML methods can capture complex, non-linear and even interactions between variables, hence leading to better predictive power in many circumstances. In view of the COVID-19 pandemic, many ML models have been developed for diagnostic or prognostic purposes. A recent review nicely summarized many of these models^4^.

Here we made of the UK Biobank (UKBB) data to build ML models to predict the severity and fatality from COVID-19, and evaluate the contributing risk factors. While predictive performance is the main concern in most previous studies, we argue that ML models can also provide important insight into individual contributing factors, and the pattern of complex relationship between risk factors and the outcome, which may not be captured by conventional linear models. We note that in the UKBB, clinical data were collected years before the outbreak of infection in 2020, which is a limitation. Ideally, the predictors should be measured at the time when the model is intended to be applied (e.g. at admission). However, we believe building ML models with previously collected clinical data is useful for several reasons.

First of all, this approach may facilitate the identification of potential causal risk factors. As the predictors are collected prior to the outbreak, there is no concern of reverse causality. In practice, infection itself will lead to changes in many clinical parameters (e.g. glucose, inflammatory markers, liver/renal functions etc.); hence it is often difficult to tell the direction of effect in cross-sectional observational studies. While many have studied risk factors on COVID-19 susceptibility or severity in the UKBB^5-7^ or other cohorts (e.g. see^4,8-11^), most relied on conventional linear models. As such, non-linear effects and interactions between variables may be missed. We hypothesize that this study will identify general or ‘baseline’ risk factors or laboratory measurements that may be predictive of outcome.

Secondly, the UKBB is a huge population-based sample (*N* close to 500,000) that enables ML models to be developed *at the general population level*. For example, at a population level, who may be more susceptible to developing severe or fatal infections? This may have implications for prioritizing individuals for specific prevention strategies (e.g. vaccination) and diagnostic testing under limited resources. Importantly, there is a relative lack of such population-level ML prediction models, and we hope this study may fill the gap.

In this study we performed four sets of analysis. In the first two sets of analysis, we built ML models to predict severity and mortality of COVID-19 within those who are tested positive for the virus. In this setting, predictive performance is of secondary concern (as predictors were not assessed at or during admission), but the predictive performance can shed light on to what extent *baseline* (pre-diagnostic) clinical characteristics contribute to severe infections. In the other two sets of analysis, we predicted severity and mortality of COVID-19 at the population level, considering subjects not known to be infected as ‘controls’. We employed XGboost, a state-of-the-art ML tool for prediction. Importantly, we also identified how different risk factors and their interactions impact on disease severity.

## Methods

### UK Biobank data

For details of UK Biobank data please refer to ref^12^. The UK Biobank is a large-scale prospective cohort comprising over 500,000 subjects aged 40–69 years when they were recruited in 2006–2010. The present analysis was conducted under project number 28732.

### COVID-19 phenotypes

COVID-19 outcome data were downloaded from data portal provided by the UKBB. Details of data release are provided at http://biobank.ndph.ox.ac.uk/showcase/exinfo.cgi?src=COVID19. Briefly, the latest COVID test results were extracted on 30 Dec 2020 (last update on 14 Dec 2020). The data also included an indicator on whether the patient was an inpatient when the specimen was taken. We consider inpatient (hospitalization) status as a proxy for severity, as more sophisticated indicators of severity cannot be reliably derived yet. Data on mortality and cause of mortality were also extracted (with latest update on 14 Dec 2020). Subjects with recorded cause of mortality “U07.1” was considered a fatal infection with laboratory-confirmed COVID-19. Please also refer to http://biobank.ndph.ox.ac.uk/showcase/exinfo.cgi?src=COVID19_tests on relevant details.

We considered a case as ‘severe COVID-19’ if the subject is an inpatient and/or if the cause of mortality is U07.1. In general we required both test result and origin to be 1 (indicating positive test and inpatient origin) to qualify as an ‘inpatient’ case. For a small number of subjects with inpatient origin=0 but result=1, and origin=1 with result=0 within 2 weeks (based on the fact that median duration of viral persistence is ∼2 weeks ^13^, we still considered those inpatient cases (i.e. assume the hospitalization was related to the infection).

For a very small number of subjects (*N*=19) whose cause of mortality was U07.1 but test result(s) was negative within one week, they were excluded from subsequent analyses.

Given that the first case of COVDI-19 in the UK was recorded on 31 Jan 2020, subjects with recorded mortality before 31 Jan 2020 (*N* = 28,930) were excluded.

### Sets of analysis

Four sets of analysis were performed. The first two sets were restricted to test-positive cases (total N=7846), with ‘severe COVID-19’ (N=2386) and death (N=477) due to COVID-19 as outcomes. Since only pre-diagnostic clinical data were available, the main objective of this analysis was to identify baseline risk factors for severe/fatal illness among the infected. We then performed another two sets of analysis with the same outcomes, but the ‘unaffected’ group was composed of the general population (N=465,728) who were *not* diagnosed to have COVID-19 or tested negative. The four sets of analysis were also referred to as cohorts A to D as shown in Table 1.In addition, we also constructed prediction models separately in males and females.

**Table 1.** The four sets of analysis performed and predictive performances (Full Model and Lite Model)

| Model | Group 1 | Group 2 | N<br>(Group 1) | N<br>(Group 2) | AUC (%) |  | 95% CI (%) |  |
| --- | --- | --- | --- | --- | --- | --- | --- | --- |
|  |  |  |  |  | Full | Lite | Full | Lite |
| A | Hospitalized or fatal cases | Non-hospitalized cases | 2,386 | 5,460 | 72.3 | 71.6 | [71.1, 73.6] | [70.3, 72.9] |
| B | Fatal cases | All other COVID-19 cases | 477 | 7,369 | 81.4 | 81.8 | [79.1, 83.8] | [79.4, 84.2] |
| C | Hospitalized or fatal cases | UKBB subjects without a COVID-19 Dx or tested -ve | 2,386 | 465,728 | 69.6 | 69.6 | [68.4, 70.8] | [68.4, 70.7] |
| D | Fatal cases | UKBB subjects without a COVID-19 Dx or tested -ve | 477 | 465,728 | 80.2 | 81.1 | [77.8, 82.6] | [78.8, 83.5] |

Dx, diagnosis. *N*, sample size. AUC was taken from the average from 5 folds of cross-validation.

### Variables included in analysis

We extracted a total of 96 clinical variables of potential relevance. Among these variables, 20 were categorical and 76 were quantitative traits. The missing rates of all variable were below 20%. We chose a relatively large set of predictors as the ML model (XGboost) is able to deal with a large number of predictors without overfitting. Another reason is that this study also aims to explore a wider variety of potential (new) risk factors for the disease. The full list of variables is shown in Table S1. Briefly, we included basic demographic variables (e.g. age, sex, ethnic group, socioeconomic status as indicated by the Townsend deprivation index), comorbidities (e.g. heart diseases, type 1 and 2 diabetes mellitus [DM], hypertension, asthma/COPD, cancer, dementia and psychiatric disorders), indicators of general health (number of medications taken, number of illnesses etc.), blood measurements (hematological measures, liver and renal function measures, metabolic parameters such as lipid levels, HbA1c etc.), anthropometric measures (e.g. waist circumference, waist-hip ratio, body mass index[BMI] etc.) and lifestyle risk factors (e.g. smoking, drinking habits etc.). For disease traits, we included information from ICD-10 diagnoses (code 41270), self-reported illnesses (code 20002) and data from follow-ups. Subjects with no records of the relevant disease from either self-reports or ICD-10 were regarded as having no history of the disease.

### Imputation

Missing values of remaining features were imputed with the R package missRanger. The program is based on missForest, which is an iterative imputation approach based on random forest (RF). It has been widely used and has been shown to produce low imputation errors and good performance in predictive models ^14^. The program missRanger is largely based on the algorithm of missForest, but uses the R package ‘ranger’ to build RFs, leading to a large improvement in speed. Predictive mean matching (pmm) was also employed to avoid imputation with values not present in the original data. We employed the default parameters (pmm.k = 3, num.trees = 100). Out-of-bag errors (in terms of classification errors or normalized root-mean-squared error) were computed which provides a guide to imputation accuracy.

### XGboost prediction model

XGboost with gradient-boosted trees was employed for building prediction models. Analysis was performed by the R package ‘xgboost’. We employed a 5-fold (nested) cross-validation strategy to develop and test the model. To avoid overoptimistic results due to choosing the best set of hyper-parameters based on test performance, the test sets were *not* involved in the tuning of hyper-parameters.

In each iteration, we divided the data into 5 folds, among which 1/5 was reserved for testing only. For the remaining 4/5 of the data, we further sampled 4/5 for training and 1/5 for hyper-parameter tuning. The best prediction model was applied to the test set. The process was repeated five times. A grid-search procedure was used to search for the best combination of hyper-parameters (e.g. tree depth, learning rate, regularization parameters for L1/L2 penalty etc.). The full range of hyper-parameters chosen or for grid-search is given in supplementary Table S2.

### Building a simplified ‘lite’ model

In addition to the full model, for easier implementation in practice, we also built a simplified ‘lite’ prediction model based on a reduced set of 27 predictors. The reduced set of variables were chosen based on the ease of being assessed or measured, which included comorbidities (see above), anthropometric measures (BMI, weight, waist circumference), demographic variables (e.g. age, sex, ethnic group) and general indicators of health (number of medications taken, number of illnesses).

### Calibration

We also assessed the calibration of the models by several measures, including Hosmer–Lemeshow test, Expected Calibration Error (ECE) and Maximum Calibration Error (MCE) ^15-18^ across 10 bins. We also attempted three approaches to further improve calibration if possible, namely Platt scaling, isontonic regression and beta calibration ^19-22^.

### Identifying and quantifying the effects of important predictors

Identification the main contributing factors to severe/fatal infection is crucial to understanding of the model and uncovering new risk factors potentially linked to the severity of infection.

In this work we primarily employed Shapley value (ShapVal)^23,24^ to assess variable importance, which is a measure based on game theory to assess the contribution of each feature. Shapley value has been shown to represent a *consistent* and locally accurate contribution of each feature ^25^. Shapley values enable local explanation of the model as they could be computed for each observation, but can also provide global importance measures. On the other hand, gain and split count may produce inconsistent estimates of global importance as shown in Lundberg et al ^25^.

Intuitively, the Shapley value of the *i*-th feature (for subject *k*) is the contribution of this feature to prediction of outcome for the individual, averaging over all possible orderings of the features (as the contribution may differ when variables enter the prediction algorithm in different orders). We ranked the global importance of features based on mean absolute ShapVal as described in previous works ^23,24^. We also attempted an alternative approach similar to ‘permutation importance’ proposed in ^26^. The method involves permuting the outcome vector to model the distribution of ShapVal under the null, and comparing the null ShapVals with the observed ShapVal. We derived a p-value as an alternative indicator of feature importance. A total of 500 permutations were performed for each model. A related index is the Shapley *interaction* value^24^, which computes the difference in Shapley value of feature *i* with and without another feature *j*. Shapley values were averaged across 5 folds. Besides, we also included the ‘gain’ measure for reference, which is the reduction of loss or impurity contributed by all splits by a specific variable.

An advantage of Shapley value is that it is calculated for each individual, so how each risk factor affect a specific person’s risk of infection/severity can be estimated as well. To illustrate this concept, we also produced decision plots for subjects at the highest, median and lowest risk of each cohort.

### Cluster analysis based on Shapley value

We also performed cluster analysis based on Shapley values to identify subgroup of patients who share similar clinical risk factors with respect to severity of infection. As introduced in (cite), this approach may be considered a form of ‘supervised’ clustering, as the outcome of interest (severe/fatal disease) is also taken into account in the clustering process. Unlike a traditional clustering approach based on the risk factors, this approach has important advantages. Firstly, the clusters derived may be more clinically relevant as the *outcome* is also considered, reducing the chance that irrelevant features contributing to the subgrouping. (An irrelevant feature will have relatively small variations in ShapVal and will not contribute substantially to the clustering). Secondly, this approach essentially considers all features on the same ‘scale’, as ShapVal are computed with respect to the outcome. Input features are often of different units and scales and but ShapVal considers feature contributions to the outcome as the unit of measure. Due to computation cost concerns, here we only perform clustering on models A (non-severe vs severe infection) and B (fatal vs non-fatal infection).

#### K-means sparse clustering

Here we performed k-means sparse clustering to uncover the underlying subgroups of the subjects based on the Shapley values of different risk factors. As the number of features included is relatively large but not all features may contribute to the underlying subgroups, we employed sparse clustering which incorporates feature selection in the clustering process. The R package “sparcl” was employed. To perform sparse k-means clustering, we need to predetermine the number of clusters and tuning parameter (L1 penalty) for feature selection^27^. The optimal number of clusters was assumed to be the same as that in a k-means clustering, which was determined by silhouette index. In this study, the range for tuning parameter (L1 bound) was firstly set to range between 2 to 6 with an interval of 0.4. Then the gap statistic^28^ was utilized to determine the optimal tuning parameter.

## Results

An overview of the sample sizes in each set of analysis is presented in Table 1. Please also refer to Table S1a and S1b for a detailed summary of case counts and covariates.

### Prediction performance

#### AUC-ROC

We performed 5-fold CV and the average AUC under the ROC curve is given in Table 1 and Table S2. Here we describe the results for the full models first. We observed better predictive performances in cohorts B (fatal cases vs outpatient cases) and D (fatal cases vs population with no known infection) when fatalities from COVID-19 were modeled. The corresponding mean AUC were 0.814 (95% CI: 0.791-0.838) and 0.802 (CI 0.778-0.826) respectively. The mean AUC for cohort A (hospitalized/fatal cases vs other cases) was 0.723 (CI: 0.711 – 0.736) and AUC for cohort C (hospitalized/fatal cases vs population with no known infection) was 0.696 (CI: 0.684 - 0.708).

As for the ‘lite’ models which include a reduced set of predictors, the predictive performances in terms of AUC are broadly similar, with estimated AUCs of 0.716, 0.818, 0.696 and 0.811 respectively.

We also conducted sex-stratified analysis (Table S2). The resulting AUC were similar to the overall analysis in males (except for model D), but were generally lower in females. This may be partially explained by lower number of severe and fatal cases in females, which leads to greater difficulty in model training.

#### Proportion of cases explained by individuals at the top k% of predicted risk

We also computed the proportion of cases explained by individuals at the highest *k*% of predicted risk (Table 2). For example, considering the full model, for prediction of mortality among infected individuals (model B), subjects at the top 5%, 10% and 15% of predicted risk explain 17.4%, 32.7% and 47.0% of total fatal cases respectively. As for prediction in the population (model D), subjects at the top 5%, 10% and 15% of predicted risk explain 23.5%, 32.9% and 36.1% of total fatal cases respectively. For prediction of severe disease among the infected (model A), subjects at the top 5%, 10% and 15% of predicted risk explain 11.2%, 21.6% and 30.7% of total cases respectively, while more than half (53.3%) of cases are explained by people at the top 30% of predicted risk. For prediction of severe case in the population, the corresponding figures were 19.7%, 29.3% and 37.1% respectively, more than half (52.8%) of cases are explained by people at the top 30% of predicted risk. Similar figures were observed for full and lite models in general (although for model D, the lite model appeared to perform better).

**Table 2.** Relative risk (RR) comparing subjects in the top and bottom *k*% of predicted risks and proportion of cases explained by those at top *k*% of predicted risk

| Full model |  |  |  |  |  | Lite model |  |  |  |
| --- | --- | --- | --- | --- | --- | --- | --- | --- | --- |
| | $k$ | Risk in top $k\%$ | Risk in bottom $k\%$ | RR | Proportion of cases explained by top $k\%$ | Risk in top $k\%$ | Risk in bottom $k\%$ | RR | Proportion of cases explained by top $k\%$ |
| Model A |  |  |  |  |  |  |  |  |  |
|  | 5 | 0.676 | 0.148 | 4.56 | 0.112 | 0.691 | 0.158 | 4.37 | 0.113 |
|  | 10 | 0.654 | 0.138 | 4.74 | 0.216 | 0.644 | 0.157 | 4.10 | 0.211 |
|  | 20 | 0.579 | 0.145 | 4.00 | 0.382 | 0.581 | 0.153 | 3.79 | 0.381 |
|  | 30 | 0.540 | 0.148 | 3.65 | 0.533 | 0.533 | 0.152 | 3.50 | 0.526 |
|  | 40 | 0.489 | 0.152 | 3.20 | 0.644 | 0.479 | 0.158 | 3.03 | 0.630 |
|  | 50 | 0.443 | 0.166 | 2.67 | 0.730 | 0.439 | 0.170 | 2.59 | 0.720 |
| Model B |  |  |  |  |  |  |  |  |  |
|  | 5 | 0.214 | 0.000 | Inf | 0.174 | 0.212 | 0.003 | 84.27 | 0.174 |
|  | 10 | 0.200 | 0.001 | 158.20 | 0.327 | 0.216 | 0.008 | 28.38 | 0.352 |
|  | 20 | 0.171 | 0.008 | 22.42 | 0.562 | 0.188 | 0.008 | 24.59 | 0.618 |
|  | 30 | 0.148 | 0.009 | 16.57 | 0.727 | 0.155 | 0.008 | 19.21 | 0.763 |
|  | 40 | 0.127 | 0.009 | 14.21 | 0.830 | 0.131 | 0.009 | 14.23 | 0.866 |
|  | 50 | 0.111 | 0.010 | 10.94 | 0.916 | 0.111 | 0.011 | 10.37 | 0.912 |
| Model C |  |  |  |  |  |  |  |  |  |
|  | 5 | 0.0201 | 0.0017 | 11.76 | 0.197 | 0.0210 | 0.0013 | 15.88 | 0.207 |
|  | 10 | 0.0149 | 0.0021 | 6.98 | 0.293 | 0.0158 | 0.0012 | 12.95 | 0.310 |
|  | 20 | 0.0109 | 0.0023 | 4.67 | 0.427 | 0.0118 | 0.0021 | 5.71 | 0.462 |
|  | 30 | 0.0090 | 0.0030 | 2.99 | 0.528 | 0.0097 | 0.0027 | 3.57 | 0.573 |
|  | 40 | 0.0075 | 0.0033 | 2.27 | 0.590 | 0.0084 | 0.0026 | 3.20 | 0.656 |
|  | 50 | 0.0069 | 0.0033 | 2.09 | 0.678 | 0.0074 | 0.0028 | 2.63 | 0.725 |
| Model D |  |  |  |  |  |  |  |  |  |
|  | 5 | 0.0048 | 0.0001 | 37.34 | 0.235 | 0.0060 | 0.0001 | 69.51 | 0.291 |
|  | 10 | 0.0034 | 0.0002 | 17.45 | 0.329 | 0.0043 | 0.0002 | 20.10 | 0.417 |
|  | 20 | 0.0019 | 0.0002 | 9.05 | 0.379 | 0.0028 | 0.0002 | 15.12 | 0.537 |
|  | 30 | 0.0019 | 0.0003 | 6.11 | 0.564 | 0.0021 | 0.0005 | 4.60 | 0.608 |
|  | 40 | 0.0018 | 0.0004 | 5.04 | 0.709 | 0.0019 | 0.0004 | 4.60 | 0.753 |
|  | 50 | 0.0016 | 0.0005 | 3.22 | 0.765 | 0.0017 | 0.0004 | 4.82 | 0.826 |

These results showed in general a strong enrichment of cases among those with high predicted risks, indicating good discriminative ability of the models and possibility to focus on highest-risk group for targeted preventions.

#### Relative risk comparing the top and bottom k%

We also computed the relative risk (RR) of infection or severe disease by comparing individuals at the top *k*% against those at the lowest *k*% of predicted risk (Table 2). For example, considering the full model, when we compared the risks of those at the top decile (top 10%) against those at the bottom decile of predicted risks, the RR was 4.74, 158.2, 6.98, 17.45 respectively for models A to D for the full model. If we compare the top 20% against the lowest 20%, the corresponding RR were 4.00, 22.42, 4.67, 9.05 respectively. The figures of RR for the lite model were similar; however we observed more extreme RR estimates for the full model under model B but the reverse under model D.

On the whole, the large RR especially for model B and D suggests that the model is able to discriminate individuals at the highest and lowest risks well.

#### Calibration

As for the calibration, please refer to Figures S6-S7. For full models, model A was well-calibrated (without using other methods for re-calibration) with ECE of 0.022 and MCE of 0.044 only. For other models, the ECE and MCE are generally larger, probably due to large difficulty in calibration with a much lower probability of the outcome. The ECEs (after re-calibration by one of the three methods) were 0.11, 0.14 and 0.10 respectively. H-L test was non-significant in models C and D. For ‘lite’ models, the ECEs were 0.017, 0.043, 0.024 and 0.199 respectively for models A to D, with non-significant H-L test results except for model B.

### Results from cluster analysis

Figure S11 shows the results based on sparse k-means clustering. We performed clustering separately in cases and controls to uncover patient subgroups with different clinical background. Here we focus on clustering results within cases. For cohort A, we found two clusters as the optimal solution. The first cluster has higher ShapVal for most risk factors, especially age, but also cnt_treatment, HbA1c, cystatin C, HDL-C and HT. ShapVal for WHR was positive for the 1st group but negative for 2nd group. The first cluster may represent a subgroup of severe cases with a larger number of clinical risk factors/comorbidities and advanced age, while the second cluster may be a distinct group with less conventional risk factors (especially obesity), yet are susceptibility to more severe infections perhaps due to other (unmeasured) factors, such as genetics.

Considering cohort B cases (fatal infections), the optimal solutions comprised three clusters. Interestingly, the 1^st^ and 3^rd^ cluster seemed to be markedly different with respect to their risk factor profiles. Mean ShapVal for age were largely negative for the 1^st^ cluster but highly positive for the other two clusters. On the other hand, mean ShapVal for waist circumference was markedly higher and positive for the 1^st^ cluster (also higher ShapVal for BMI, WC). The 3^rd^ cluster was characterized by the highest mean ShapVal for age, and higher (positive) ShapVal for mainly cnt_treatment, HbA1c and T2DM. The results suggest that there may exist patho-physiologically distinct subgroups of patients who have had a fatal infection. The 1^st^ cluster represents a subgroup with younger age but with higher BMI and proportion of obesity. The 3^rd^ cluster represented another group with advanced age, more comorbidities and higher proportion of glucose abnormalities or DM. The 2^nd^ cluster was in between.

### Important contributing variables identified

Here we primarily report the results of the full model as a more complete set of predictors is included. The Shapley dependence plots (ranked by mean absolute ShapVal) of the top 15 features (full model) are shown in Figure 1 and those of top 6 features for lite model are in Figure 2. For more complete plots (up to 30 variables) with ranking by mean abs(ShapVal) or p-values, please refer to Figures S1-4.

**Figure 1.**
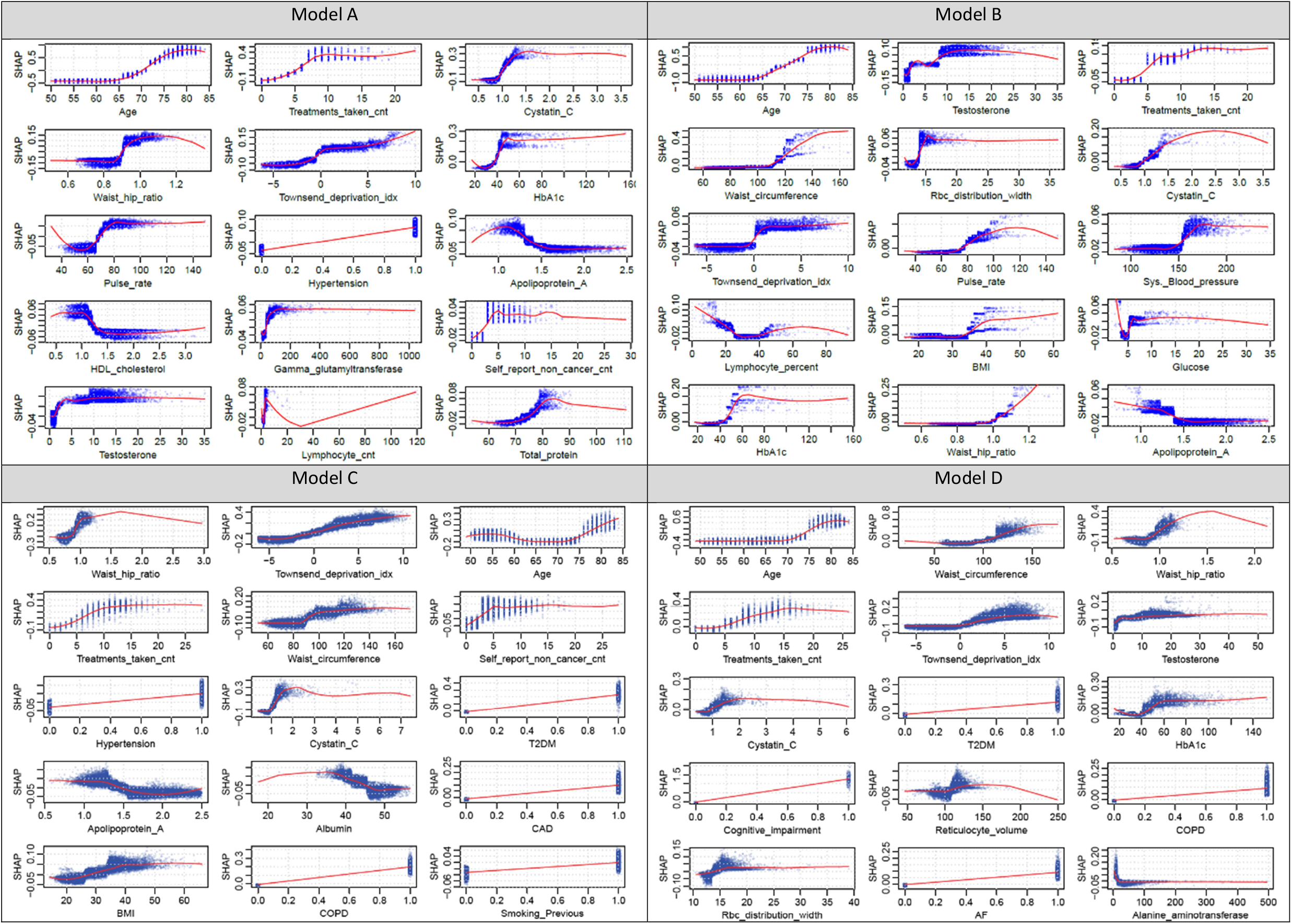
Shapley value dependence plots of top 15 risk factors ranked by mean abs(shapley value) (Full model) for Models A, B, C, D respectively.

**Figure 2.**
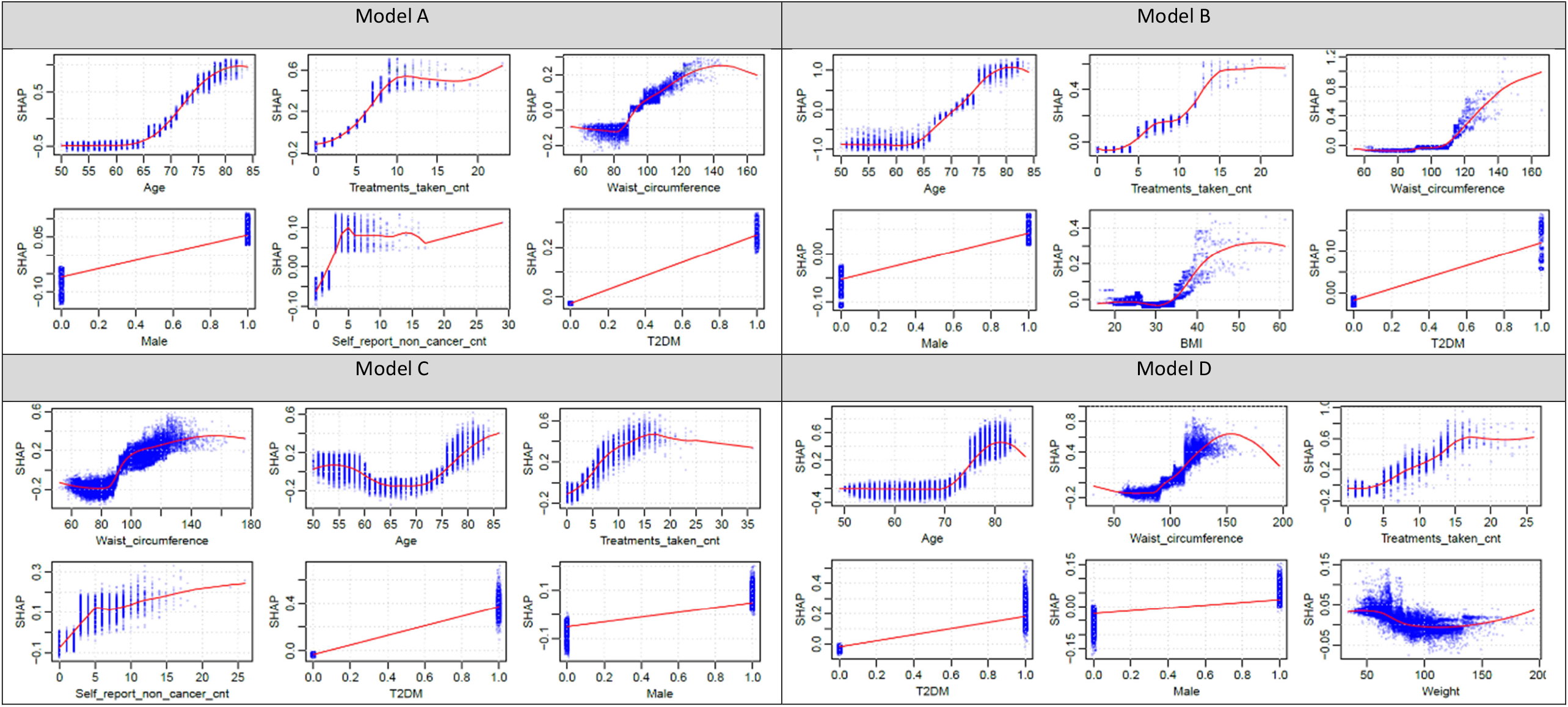
ShapVal dependence plots of top 6 risk factors ranked by mean abs(shapley value) (Lite model) for Models A, B, C, D respectively.

Full ShapVal analysis results on all variables are given in Tables S3a-c. Top 10 variables (ranked by either ShpaVal or p-value) from the full model are presented in Tables 3 and 4 while top 5 from the lite model are presented in Tables 5-6. We also included variable importance by gain and plots are presented in Figure S5.

**Table 3.** Top 10 risk factors ranked by mean absolute Shapley value for Model A, B, C and D (Full Model)

| Cohort A |  |  | Cohort B |  |  |
| --- | --- | --- | --- | --- | --- |
| Risk factor | ShapVal | P-Val | Risk factor | ShapVal | P-val |
| Age | 0.442 | 0.002 | Age | 0.708 | 0.002 |
| Treatments taken cnt | 0.093 | 0.002 | Testosterone | 0.069 | 0.002 |
| Cystatin C | 0.088 | 0.002 | Treatments taken cnt | 0.048 | 0.002 |
| Waist hip ratio | 0.085 | 0.002 | Waist circumference | 0.035 | 0.002 |
| Townsend deprivation idx | 0.059 | 0.004 | Rbc distribution width | 0.027 | 0.002 |
| HbA1c | 0.056 | 0.002 | Cystatin C | 0.024 | 0.002 |
| Pulse rate | 0.048 | 0.002 | Townsend deprivation idx | 0.023 | 0.002 |
| Hypertension | 0.048 | 0.002 | Pulse rate | 0.019 | 0.004 |
| Apolipoprotein A | 0.027 | 0.016 | Sys. Blood pressure | 0.016 | 0.002 |
| HDL cholesterol | 0.026 | 0.016 | Lymphocyte percent | 0.015 | 0.004 |
| Cohort C |  |  | Cohort D |  |  |
| Risk factor | ShapVal | P-val | Risk factor | ShapVal | P-val |
| Waist hip ratio | 0.113 | 0.002 | Age | 0.243 | 0.002 |
| Townsend deprivation idx | 0.096 | 0.002 | Waist circumference | 0.049 | 0.008 |
| Age | 0.088 | 0.002 | Waist hip ratio | 0.046 | 0.044 |
| Treatments taken cnt | 0.063 | 0.002 | Treatments taken cnt | 0.030 | 0.026 |
| Waist circumference | 0.044 | 0.002 | Townsend deprivation idx | 0.030 | 0.120 |
| Self-report non-cancer cnt | 0.043 | 0.002 | Testosterone | 0.028 | 0.096 |
| Hypertension | 0.036 | 0.002 | Cystatin C | 0.017 | 0.178 |
| Cystatin C | 0.030 | 0.024 | T2DM | 0.016 | 0.004 |
| T2DM | 0.030 | 0.002 | HbA1c | 0.012 | 0.218 |
| Apolipoprotein A | 0.024 | 0.052 | Cognitive impairment | 0.010 | 0.002 |

**Table 4.** Top 10 risk factors ranked by P-value, listing only factors which are *not* yet included in Table 3 for Model A, B, C and D (Full Model).

| Cohort A |  |  | Cohort B |  |  |
| --- | --- | --- | --- | --- | --- |
| Risk factor | P-val | ShapVal | Risk factor | P-val | ShapVal |
| T2DM | 0.004 | 0.010 | BMI | 0.002 | 0.015 |
| Self-report non-cancer cnt | 0.008 | 0.018 | Glucose | 0.002 | 0.015 |
| Depression | 0.008 | 0.004 | HbA1c | 0.002 | 0.014 |
| CAD | 0.016 | 0.002 | Weight | 0.002 | 0.010 |
| Cancer diagnosed by doctor | 0.026 | 0.000 | Mean platelet volume | 0.002 | 0.009 |
| Alcohol intake (Occasions) | 0.028 | 0.002 | T2DM | 0.002 | 0.007 |
| AF | 0.028 | 0.000 | Sleep Duration | 0.002 | 0.006 |
| Smoking (Current) | 0.036 | 0.000 | T1DM | 0.002 | 0.003 |
| Gamma glutamyltransferase | 0.046 | 0.021 | Cognitive impairment | 0.002 | 0.003 |
| Wbc cnt | 0.046 | 0.014 | CAD | 0.002 | 0.003 |
| Cohort C |  |  | Cohort D |  |  |
| Risk factor | P-val | ShapVal | Risk factor | P-val | ShapVal |
| COPD | 0.002 | 0.015 | COPD | 0.008 | 0.008 |
| Depression | 0.002 | 0.009 | AF | 0.010 | 0.007 |
| Cognitive impairment | 0.002 | 0.007 | Bipolar | 0.010 | 0.001 |
| CAD | 0.004 | 0.017 | Heart failure | 0.012 | 0.003 |
| Ethnic (Asian/Asian British) | 0.004 | 0.007 | Ethnic (Black/Black British) | 0.020 | 0.002 |
| Heart failure | 0.004 | 0.007 | T1DM | 0.020 | 0.001 |
| AF | 0.004 | 0.006 | Haemorr stroke | 0.024 | 0.002 |
| Smoking (Previous) | 0.006 | 0.015 | CAD | 0.028 | 0.005 |
| Haemorr stroke | 0.012 | 0.001 | Hypertension | 0.034 | 0.005 |
| Ethnic (Black/Black British) | 0.020 | 0.001 | Smoking (Current) | 0.050 | 0.002 |

**Table 5.** Top 5 risk factors ranked by mean absolute Shapley value for Model A, B, C and D (Lite Model)

| Cohort A |  |  | Cohort B |  |  |
| --- | --- | --- | --- | --- | --- |
| Risk factor | ShapVal | P-Val | Risk factor | ShapVal | P-val |
| Age | 0.496 | 0.002 | Age | 0.721 | 0.002 |
| Treatments taken cnt | 0.121 | 0.002 | Treatments taken cnt | 0.079 | 0.014 |
| Waist circumference | 0.085 | 0.002 | Waist circumference | 0.071 | 0.040 |
| Male | 0.058 | 0.002 | Male | 0.048 | 0.010 |
| Self-report non-cancer cnt | 0.054 | 0.004 | BMI | 0.034 | 0.242 |
| Cohort C |  |  | Cohort D |  |  |
| Risk factor | ShapVal | P-val | Risk factor | ShapVal | P-val |
| Waist circumference | 0.153 | 0.002 | Age | 0.226 | 0.002 |
| Age | 0.12 | 0.002 | Waist circumference | 0.106 | 0.086 |
| Treatments taken cnt | 0.102 | 0.002 | Treatments taken cnt | 0.057 | 0.114 |
| Self-report non-cancer cnt | 0.064 | 0.002 | T2DM | 0.026 | 0.014 |
| T2DM | 0.05 | 0.002 | Male | 0.025 | 0.074 |

**Table 6.** Top 5 risk factors ranked by P-value, listing only factors which are *not* yet included in Table 5 for Model A, B, C and D (Lite Model).

| Cohort A |  |  | Cohort B |  |  |
| --- | --- | --- | --- | --- | --- |
| Risk factor | P-val | ShapVal | Risk factor | P-val | ShapVal |
| T2DM | 0.047 | 0.002 | T2DM | 0.027 | 0.006 |
| Smoking (Current) | 0.026 | 0.004 | Cognitive impairment | 0.015 | 0.006 |
| Depression | 0.015 | 0.016 | T1DM | 0.009 | 0.020 |
| Alcohol drinker (Current) | 0.013 | 0.020 | Bipolar | 0.006 | 0.024 |
| CAD | 0.010 | 0.022 | AF | 0.011 | 0.036 |
| Cohort C |  |  | Cohort D |  |  |
| Risk factor | P-val | ShapVal | Risk factor | P-val | ShapVal |
| COPD | 0.024 | 0.002 | Cognitive impairment | 0.012 | 0.002 |
| Ethnic (Asian/Asian British) | 0.016 | 0.002 | Bipolar | 0.002 | 0.018 |
| Cognitive impairment | 0.008 | 0.002 | COPD | 0.014 | 0.032 |
| Male | 0.049 | 0.004 | AF | 0.013 | 0.044 |
| CAD | 0.023 | 0.004 | Heart failure | 0.004 | 0.046 |

As for interaction analysis, top results are presented in Table 7 and full results in Tables S4-5. Plots are presented in Figure 3 (top two interacting pairs from each model) and Figures S8-9 (top 6 interacting pairs).

**Table 7.** Top interacting pairs of variables ranked by ShapVal (Full model)

| Cohort A |  |  | CohortB |  |  |
| --- | --- | --- | --- | --- | --- |
| Risk factor 1 | Risk factor 2 | Value | Risk factor 1 | Risk factor 2 | Value |
| Waist hip ratio | Age | 150 | Testosterone | Age | 195 |
| Treatments taken cnt | Age | 149 | Waist circumference | Age | 95 |
| HDL cholesterol | Age | 86 | BMI | Age | 81 |
| Age | Hypertension | 85 | Treatments taken cnt | Age | 63 |
| Cystatin C | Age | 84 | Pulse rate | Age | 57 |
| Cohort C |  |  | Cohort D |  |  |
| Risk factor 1 | Risk factor 2 | Value | Risk factor 1 | Risk factor 2 | Value |
| Waist hip ratio | Age | 709 | Waist hip ratio | Age | 893 |
| Waist hip ratio | Treatments taken cnt | 494 | Testosterone | Age | 832 |
| Townsend deprivation idx | Treatments taken cnt | 481 | Waist circumference | Age | 626 |
| Townsend deprivation idx | Waist hip ratio | 450 | Townsend deprivation idx | Age | 565 |
| Albumin | Waist hip ratio | 407 | Treatments taken cnt | Age | 405 |

**Figure 3.**
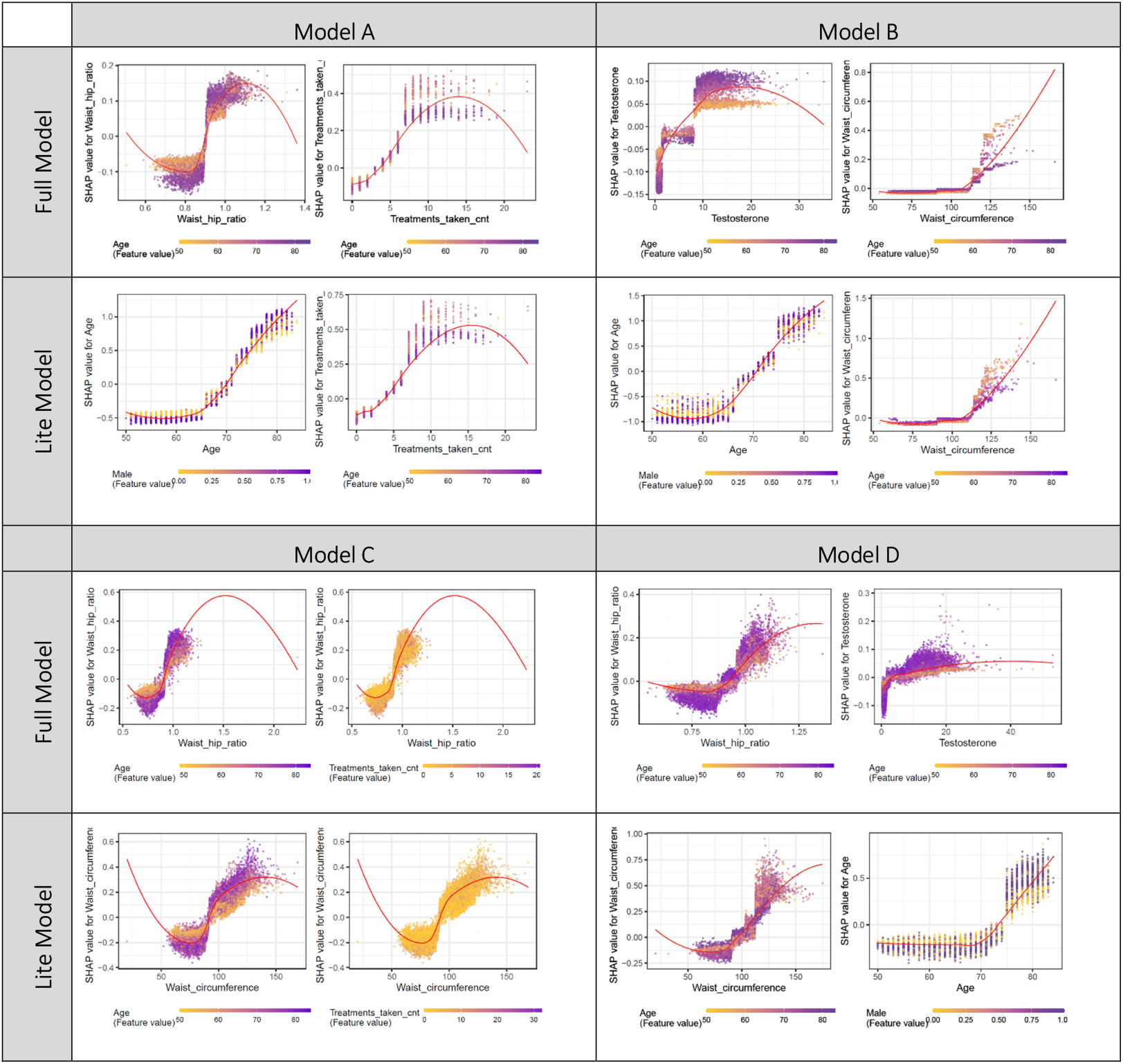
ShapVal interaction plots of the Full and Lite model for the top 2 interaction values of Model A, B, C, D respectively.

#### Model A (hospitalized/fatal cases vs outpatient cases)

For this set of analysis, the top 5 contributing features by ShapVal included age, number of medications received (cnt_treatment), cystatin C, Townsend Deprivation index (TDI), waist-hip ratio (WHR), followed by HbA1c. Higher levels of these risk factors generally lead to higher severity of disease among the infected. Interestingly, Shapley dependence plots revealed potential *non-linear* effects of risk factors on the outcome. For example, age of ∼65 or above was associated with a marked increased risk of severe/fatal infection. Elevated risks were also observed true for HbA1c > ∼40 mmol/mol and number of drugs received >= ∼5. Impaired renal function (raised cystatin C above ∼1 mg/L) was also linked to worse outcomes. For WHR, levels of ∼0.9 or higher appeared to be associated with marked increase in risks. For the effect of other features please also refer to Figure 1. We note that at extreme ends of the variables, the observations are sparse so the trend shown by the loess curve may not be reliable (this also applies to other cohorts). Variable importance based on gain revealed similar patterns of important features (Figure S5).

If we consider the ‘p-value’ or permutation importance (PermImp) measure, variables with top 10 |ShapVal| also have significant p-values. Type 2 diabetes were among the top 10 by PermImp but not |ShapVal|. Depression and CAD also have low p-values (<0.02) but not listed among top 30 by ShapVal.

With regards to interactions between variables, most the top pairs of interacting variable involve age (Figure 3, Tables S4-5). For example, younger individuals were observed to have more extreme ShapVal at similar ranges of cnt_treatment (number of medications). The effect of WHR on severity was more marked among the elderly, and the same was true for HDL cholesterol (low HDL is a risk factor).

#### Model B (fatal cases vs outpatient cases)

The top 5 contributing variables by ShapVal included age, testosterone, cnt_treatment, waist circumference (WC) and red cell distribution width (RDW), which were followed by cystatin C, TDI pulse rate, systolic blood pressure (SBP) and percentage of lymphocytes.

Again certain non-linear ‘threshold’ effects appeared to be present for many top-ranked features. For age, the risk for mortality was more marked beyond ∼65. Higher levels of all the above risk factors (RFs) (except percentage of lymphocytes which showed a U-shaped relationship) were associated with higher mortality, but the effects were non-linear and showed threshold effects. Regarding top results using PermImp, 8 out of 10 predictors ranked high by |ShapVal| also had the lowest p-values (lowest p-value = 0.002 since we performed 500 permutations). Quite a number of covariates ranked high with p-value (PermImp)=0.002, including HbA1c, type 1 and type 2 DM, weight, mean platelet volume etc.

Variable importance based on gain yielded similar result (Figure S5). As for interaction between the variables, again the interactions were most prominent with age. For example, the effects of waist circumference and BMI (when exceeding a threshold of around 110 cm and 35 kg/m^2^) on mortality were more prominent among younger individuals. The effects of testosterone and HbA1c however were more marked in older subjects.

#### Model C (hospitalized/fatal cases vs population with no known infection)

Based on ShapVal, WHR was the top contributing variable and WC was ranked 5^th^, suggesting central obesity may be a stronger predictor for severe disease than BMI alone (BMI was ranked 13^th^ by ShapVal). Similar to before, TDI and age were ranked among the top. For age, slightly unexpectedly, a U-shaped curve was observed, which suggests lowest risk at the age group of ∼65-70. Note that model C may also capture RFs related to susceptibility to infection. It is possible, for instance, that younger subjects had higher risks of exposure due to work or social interactions. Among the top 10, two are related to multi-comorbidities in general (cnt_treatment/cnt_noncancer). Increased cystatin C and lower apoplipoprotien A were also associated with higher susceptibility to severe infections, and HT and Type 2 DM were also among the top 10. When considering PermImp as the ranking criteria, COPD and depression were also ranked among the top 10 though not top-listed by ShapVal.

Interaction plot (Figure 3) shows WHR may interact with age, with elderly individuals showing more prominent effects from changes in WHR.

#### Model D (fatal cases vs population with no known infection)

Age was the top feature, followed by WC, WHR, number of drugs taken and TDI. Other top features included testosterone, cystatin C, Type 2 DM, HbA1c and dementia. An increase of these features and the mentioned disorders generally lead to increased risk of mortality.

Shapley interaction analysis suggested the top interacting pairs involved age and some of top contributing features. The effects of WHR, testosterone and TDI were more prominent among the elderly. Another noteworthy interaction is between type 2 DM and age. The increased risk of mortality with increasing age is more marked in type 2 diabetic patients.

As for important variables from sex-stratified analysis, the top variables were similar which included e.g. age, WC/WHR, cystatin C, HbA1c, number of medications received, socioeconomic status (as reflected by TDI), among others (Table S3c).

### Results from Lite model

Here we highlight top contributing features for the ‘lite’ models consisting of 27 predictors. Remarkably, the top 3 features (ranked by ShapVal) were consistent across all four models. These features included age, cnt_treatment received and WC (WHR was not included in lite model as WC is easier to measure). Of note, sex and Type 2 DM were ranked among the top 6 across all models.

If we consider PermImp as the ranking criteria (further ranked by ShapVal if PermImp is equal) and the top 5 predictors, age, cnt_treatment and WC are still highly ranked and top-listed in at least 2 models. Notably, Type 2 DM was ranked among top 5 in all models, while dementia was ranked 2^nd^ in models B and D just after age.

### Individual Shapley decision plots and online calculator

Note that we also showed individual Shapley decision plot for three subjects with the highest, median and lowest predicted risks in each cohort (Fig S10). The y-axis is based on a log-odds scale.

To facilitate further research and studies on risk prediction models, we also constructed an online risk calculation tool at http://labsocuhk.ddns.net:8889/covid19/. The online tool can also construct a Shapley decision plot based on individual risk factors.

## Discussions

In this study we performed four sets of analysis, predicting severe or fatal COVID-19 cases among the affected individuals or in the population. We also identified risk factors for increased severity or mortality from infection, which may shed light on disease mechanisms.

### Prediction of severity/mortality

In general, our prediction models achieved reasonably good predictive power. The models predicted mortality (AUC ∼81-82%) better than severity of disease. As discussed earlier, in the absence of better alternatives, hospitalization (test performed as inpatient) was used as a proxy for severity. However, reasons or criteria for hospitalization may vary across individuals or hospitals, and some tests may be performed in in-patients for surveillance or due to other confirmed/suspected cases in the ward. On the other hand, mortality from infection is a more objective outcome.

By assessing the proportion of cases explained by those at the top k% of predicted risks, we observed in general a strong enrichment of cases among those with high predicted risks, indicating good discriminative ability of the models and suggesting the possibility to focus on highest-risk group for targeted preventive measures or treatment. Similar strong enrichment was also observed for the lite model with fewer predictors. We also observed large relative risks when comparing subjects at the high percentiles against those at lower percentiles. For example, for the prediction of mortality among the infected, the RR was up to 158 times (∼20% vs 0.1%) when comparing and top and bottom deciles using the full model, and 28.38 times when considering the simplified model (∼21% vs 0.8%). This suggests the model may potentially be used to stratify patients into different levels of risks and to prioritize those who may have higher risk of deterioration, who may need early medical attention or admission. As our model only relies on demographic data and information on comorbidities (especially for the lite model), risk stratification may be done even at the start of the illness without other blood or imaging results.

### Prediction of severity/mortality

Regarding the predictive performance, the models predicted mortality (AUC ∼81-82%) better than severity of disease. As discussed earlier, in the absence of better alternatives, hospitalization (test performed as inpatient) was used as a proxy for severity. However, reasons or criteria for hospitalization may vary across individuals or hospitals, and some tests may be performed in in-patients for surveillance or due to other confirmed/suspected cases in the ward. On the other hand, mortality from infection is a more objective outcome.

A number of studies focused on prediction of severity/mortality of disease (corresponding to our prediction in cohorts A and B) were available and reviewed in ref ^4^. For model A (prediction of severity among infected), the AUC is 72.3%, which is moderate but not as good as many previous ML models for severity prediction^4^. The AUC for prediction of mortality is much higher (AUC = 81.4%), although we noted some studies have reported higher predictive power from clinical symptoms, blood biochemistry on admission and imaging features^4^. We understand that without access to the above features, predictive performance may be inferior. On the other hand, due to heterogeneity of clinical samples, treatment approaches, model evaluation methods and other features across studies, direct comparisons of predictive performance across studies may be difficult. Here we are not aimed at deriving a highly accurate prediction model; the main purpose is to identify general or ‘baseline’ risk factors for severe disease, thereby gaining insight into disease pathophysiology. However, we also showed that such clinical features or blood measurements, even when collected much earlier in time, may still be quite highly predictive of outcomes and hence may be incorporated into existing prediction algorithms. The models here may also be useful when blood results or imaging are not available (e.g. before admission) and the goal is to quickly classify a patient’s risk.

For cohorts C and D, the general population (with no known infection) was also included. Compared to cohorts A and B, the identified risk factors may also increase the overall susceptibility to infection. The AUC for cohort C (severe/fatal disease) is ∼70% but is much higher when mortality is considered as the outcome (AUC∼80%). To our knowledge, there are still very few predictive models built at a *general population level* to identify susceptible individuals; this work is among the first to employ an ML approach to risk prediction of COVID-19/severe infection at a population level. DeCaprio et al.^29^ proposed an ML model to assess the vulnerability to COVID-19 in the population. However, due to limited data, no actual COVID-19 patients were included and ‘proxy’ outcomes were used instead. Models were built from mainly demographic and comorbidity data to predict hospitalization due to acute respiratory distress syndrome, pneumonia, influenza, acute bronchitis and other respiratory tract infections.

Another very recent study (‘QCOVID’ study) from the UK^30^ utilized general practice records from 6.08 million adults (age 19 to 100) as derivation cohort and 2.17 million adults as the validation set. Mortality from COVID-19 was the primary outcome and a survival model (sub-distribution hazard model)^31^ was used to predict mortalities. The predictors included demographic (e.g. age, TDI, ethnicity), lifestyle (e.g. BMI, smoking) and a large range of comorbid conditions. The resulting Harrell’s C (comparable to AUC) was 0.928. However, we note that the QCOVID study includes subjects at a much younger age range (19 or above), which will improve predictive performance as age is by far the most important predictor of mortality with markedly reduced risk in younger subjects. For example, if we refer to age-specific predictive performance (Supplementary Table C of the paper), Harrell’s C for mortality was 0.678, 0.831, 0.812 and 0.814 in 50-59, 60-69, 70-79 and 80+ year olds respectively, for males in the first follow-up period (24 Jan to 30 Apr 2020). For females, the corresponding numbers were 0.618, 0.77, 0.866 and 0.821. These numbers reflect lower predictive power when restricted to a narrower age range. One main difference between the present work and the above study is that we employed a *machine learning* approach which is able to capture also non-linear and more complex interaction effects. As shown in our Shapley dependence plots, the models were able to reveal non-linear effects in a data-driven manner. We also included a number of blood measurements to shed light on potential new risk factors and mechanisms underlying the disease. The QCOVID study employed a survival model (sub-distribution hazard) that accounts for time-to-event and competing risks; however, the proportional hazards assumption is required which may not hold due to restrictions/interventions introduced during the period (i.e. time-dependent associations may be present).

A few other studies have studied risk factors (especially comorbidities) for COVID-19 infection in the UKBB. For example, Atkins et al. ^5^ studied elderly subjects (age>65) in UKBB, and found that hypertension, history of falls, coronary heart disease and type 2 diabetes and asthma as the top comorbidities among those hospitalized cases. The analysis was restricted to the elderly population however. In a more recent work, McQueenie et al. ^6^ studied multi-comorbidity and polypharmacy on the risk of developing the disease. The main risk factors were having >=2 long-term conditions, cardiometabolic disorders and polypharmacy were associated with heightened risk of infection. Among individuals with multi-comorbidities, severe obesity and impaired renal function may lead to increased risks. Another study of primary care patients in the UK revealed deprivation, males, older age, ethnicity (being black) and chronic renal disease were associated with higher risks of being tested positive. Another large-scale British primary care study of more than 17 million subjects revealed similar risk factors as above ^32^. There is also a relatively large literature on the study of risk factors associated with severe or fatal disease^8-11,33-36^. Some commonly reported risk factors included age, sex, obesity, diabetes, hypertension, cardiometabolic and respiratory disorders. As discussed above, an important difference between the above epidemiological studies and the current work is that we employed a machine learning approach (XGboost), which may capture non-linear and interaction effects, while other studies mostly used regression models that assumes linear and additive effects of covariates. We also performed a comprehensive analysis including four models covering different outcomes and both infected and population cohorts.

### Highlights of potential risk factors

For limit of space, we shall only highlight the top 5-10 risk factors ranked by Shapley values here. Across the four cohorts, age and cardiometabolic risk factors predominate the top risk factors. Age and WHR/waist circumference was ranked among top 5 across all four cohorts. The number of medications of taken was in top 5 across all cohorts. Of note, cystatin C (reflecting renal function) was among the top 10 in all cohorts. HbA1c was a top-10 risk factor across two cohorts (A/D), and type 2 DM was also among the top 10 for cohorts C and D. Townsend deprivation index (reflecting socioeconomic status) was also ranked among the top 10 in most models. As described above, results from ‘lite’ model were generally in line with those from the full model, with age, WC and cnt_treatment consistently ranked as the top 3.

Obesity has been observed to be a major risk factor for susceptibility or severity of infection in the UK Biobank ^7,37^, and in many other studies^38,39^. The observation that waist circumference/WHR were highly ranked suggests that *central* obesity is a major risk factor and may be a better predictor of severity than BMI alone.

Among other comorbidities, another major risk factor we identified was impaired renal function (IRF), as reflected by elevated risks with raised urea and cystatin C. Several studies also suggested IRF increases risk of mortality^36,40,41^, although it is probably not as widely recognized as cardiometabolic disorders as a main risk factor. Since COVID-19 itself may lead to renal failure, our findings specifically suggest that underlying or baseline IRF is an important risk factor. The high ranking of cystatin C also indicates this measure may better reflect renal function than urea or creatinine (which were also included in our analysis) ^42,43^, and may serve as a superior predictor for COVID-19 severity. HbA1c reflects glycemic control, and it is unsurprising that it showed up as a risk factor for severity or mortality. Diabetes has been shown to raise the risk and severity of infection^44,45^.

Other potential risk factors briefly highlighted below were less reported; as most were listed only once or twice among the top 10 list, and for some their ShapVal were close to other risk factors, further replications are required. For example, testosterone was top-ranked by XGboost (for mortality), with higher levels associated with increased risk. This may partially reflect that males are at a higher risk of fatal infections, but it remains to be studied whether testosterone itself is involved in the pathophysiology of severe COVID-19, as the ML model chose this variable instead of sex. Studies have suggested elevated or reduced testosterone levels may be both associated with a more severe clinical course^46^. Also, interestingly, 5-alpha-reductase inhibitors or androgen-deprivation therapy have been shown be associated with lower risk or severity of disease ^47,48^. We also found a few hematological indices that may be potential risk factors. High red cell distribution width (RDW) was associated with mortality in our study and was also identified in a recent meta-analysis of three studies as a risk factor ^49^. Low lymphocyte percentage was a top-10 risk factor in cohort B, which may be related to immune functioning and response to infections. Lymphopenia has been reported as a main hematological finding in those with severe illness ^18,50^. Most previous studies considered hematological indices at admission or during hospitalization. Slightly surprisingly, this study suggested that high RDW or reduced lymphocyte percentage *prior to the diagnosis* may also be predictive of worse outcomes.

Among the diseases being included as covariates, Type 2 DM is most consistently ranked among the top, no matter in the full or lite models, and regardless of ranking by ShapVal or PermImp (p-value). We noted some discrepancy between the ranked results based on ShapVal and those based on PermImp. In general, the latter measure favors binary variable while ShapVal alone tends to rank continuous variables higher. We are unsure about the exact reason for this discrepancy but it may be an interesting topic for further methodology studies. If we employed a composite ranking criteria based on PermImp then by ShapVal (if equal PermImp), then a few more diseases were ranked among the top 10, such as hypertension and COPD. For model D, dementia, type 1 and type 2 DM, COPD, atrial fibrillation (AF), bipolar disorder and heart failure were also top-ranked, suggesting that a range of chronic cardiovascular, respiratory and neuropsychiatric conditions may be associated with increased mortality.

### Full and lite prediction models

We note that the simplified (‘lite’) prediction model has very similar predictive performance (as assessed by AUC) to the ‘full’ model with a larger panel of predictors. However, it is important to note that features associated with the outcome may not always improve predictive power. AUC is relatively insensitive to detecting changes in predictive performance when additional risk factors are added ^51-53^.

For example, Pencina et al. ^51^ showed that in the prediction of CVD risk in the Women’s Health Study, adding extra established risk factors often result in minimal improvements in AUC. For instance, in a model with age, SBP and smoking, adding any lipid measures result in only an increase of 0.01 in AUC from the baseline of 0.76. In the same vein, starting from a fullprediction model [containing Ln(age), Ln(SBP), smoking, Ln(TC), Ln(HDL)], deleting any one of these established CVD risk factors (except age) results in a very small reduction of AUC of <0.02. In general, for a model with high baseline AUC from existing predictors (e.g. age, sex and obesity in the case of COVID-19), including additional predictors may not result in much improvement in discriminative power or AUC. ^54^

Nevertheless, it is still much valuable to study variable importance (e.g. ShapVal) from ML model as they may shed light on the pathophysiology of the disease. For example, many factors such as age and diabetes may lead to poorer renal function (and higher cystatin C), which in turn may increase the severity of infection. Given that age, diabetes and other main comorbidities are already modeled, adding cystatin C may not improve discriminative power of our model. However, its inclusion may still change the predicted probability of outcome, which will also be reflected in Shapley values. This example illustrates that exploring features with high VarImp may have the potential in improving our understanding of disease mechanisms.

### Limitations

Some limitations have been discussed above, for example the use of hospitalization as a proxy for severity, and that the predictors were recorded prior to the pandemic. We briefly discuss other limitations here. The UK biobank is a very large-scale study with detailed phenotypic data, but still the number of fatal cases is relatively small. Also, UKBB is not entirely representative of the UK population, as participants tend to be healthier and wealthier overall^55^. Also, it remains to be studied whether the findings are generalizable to other populations. Symptom measures and lung imaging features were not available. Despite adjusting for a rich set of predictors and that all were recorded prior to the outbreak, causality cannot be confirmed from the current study, as there is risk of residual confounding by unknown factors. The current study was performed on a cohort with age>50 and may not predict well in younger individuals. In cohorts C and D, the population with no known infection was regarded as controls. It is expected that some may become infected in the future, and some may have been infected but not tested; however, the chance of missing cases of severe infection is probably not high. Since the UKBB represents a relatively healthy population with low rate of severe COVID-19 cases so far (∼0.5%), we expect the use of ‘unscreened’ controls is unlikely to result in substantial bias.

Regarding the ML model, XGboost is a state-of-the-art method that has been consistently shown to the best or one of the best ML methods in supervised learning tasks/competitions ^56^ (especially for tasks not involving computer vision or natural language processing). Nevertheless, other ML methods may still be useful or may uncover novel risk factors. Assessing variable importance is a long-standing problem in ML; here we mainly employed Shapley values which is both computationally fast and was shown to have good theoretical properties^23,24^.

## Conclusions

In conclusion, we identified a number of baseline risk factors for severe/fatal infection by an ML approach. The Shapley plots revealed possible non-linear and ‘threshold’ effects on risks of severity. To summarize, age, central obesity, impaired renal function, multi-comorbidities, cardiometabolic abnormalities or disorders (especially DM) and low socioeconomic status may predispose to poor outcomes, among other risk factors. The prediction models (for cohorts C/D) may be useful at a population level to identify those susceptible to developing severe/fatal infections, facilitating targeted prevention strategies. Further replication and validation in independent cohorts are required to confirm our findings.

## Data Availability

The UK Biobank data is available to registered researchers.

## Supplementary Materials are available at

https://drive.google.com/drive/folders/17O1PUd1tE5gk_lFiXUyrn2vz7KUb_fRM?usp=sharing An online risk calculation tool is available at http://labsocuhk.ddns.net:8889/covid19/.

## Author Contributions

Conception and design: HCS. Analytic methodology: HCS and KCYW. Data analysis: KCYW (main), YX, HCS. Interpretation: HCS and KCYW. Supervision of study: HCS. Drafting of manuscript: HCS, with input from KCYW.

## Acknowledgements

This work was supported partially by the Lo Kwee Seong Biomedical Research Fund from The Chinese University of Hong Kong. We thank Prof. Pak Sham for support on data access and analyses.

## Conflicts of interest

The authors declare no conflict of interest.

## Notes

### Competing Interest Statement

The authors have declared no competing interest.

### Author Declarations

The UK Biobank study has received ethical approval from the NHS National Research Ethics Service North West (16/NW/0274).

